# Malaria species positivity rates among symptomatic individuals across regions of differing transmission intensities in Mainland Tanzania

**DOI:** 10.1101/2023.09.19.23295562

**Authors:** Zachary R. Popkin Hall, Misago D. Seth, Rashid A. Madebe, Rule Budodo, Catherine Bakari, Filbert Francis, Dativa Pereus, David J. Giesbrecht, Celine I. Mandara, Daniel Mbwambo, Sijenunu Aaron, Samwel Lazaro, Jeffrey A. Bailey, Jonathan J. Juliano, Deus S. Ishengoma

## Abstract

Recent data indicate that non-*Plasmodium falciparum* species may be more prevalent than previously realized in sub-Saharan Africa, the region where 95% of the world’s malaria cases occur. Although *Plasmodium malariae, Plasmodium ovale* spp., and *Plasmodium vivax* are generally less severe than *P. falciparum*, treatment and control are more challenging, and their geographic distributions are not well characterized. In order to characterize the distribution of malaria species in Mainland Tanzania (which has a high burden and geographically heterogeneous transmission levels), we randomly selected 3,284 samples from 12,845 samples to determine presence and parasitemia of different malaria species. The samples were collected from cross-sectional surveys in 100 health facilities across ten regions and analyzed via quantitative real-time PCR to characterize regional positivity rates for each species. *P. falciparum* was most prevalent, but *P. malariae* and *P. ovale* were found in all regions except Dar es Salaam, with high levels (>5%) of *P. ovale* in seven regions (70%). The highest positivity rate of *P. malariae* was 4.5% in Mara region and eight regions (80%) had positivity rates ≥1%. We also detected three *P. vivax* infections in the very low-transmission Kilimanjaro region. While most samples that tested positive for non-falciparum malaria were co-infected with *P. falciparum*, 23.6% (n = 13/55) of *P. malariae* and 14.7% (n = 24/163) of *P. ovale* spp. samples were mono-infections. *P. falciparum* remains by far the largest threat, but our data indicate that malaria elimination efforts in Tanzania will require increased surveillance and improved understanding of the biology of non-falciparum species.

## Introduction

Sub-Saharan Africa (SSA) accounted for 95% of malaria cases and 96% of malaria deaths in 2021[1]. Although *Plasmodium falciparum* is the most prevalent and deadliest human malaria species in the SSA region, four other species are present (*P. vivax, P. malariae, P. ovale curtisi*, and *P. ovale wallikeri*). Furthermore, recent research suggests that these four species are more prevalent than previously thought and their prevalence may increase in the context of intensive *P. falciparum* control/elimination[2–8]. This possibility is particularly salient in light of current malaria elimination goals stemming from the WHO Global Technical Strategy[9], which aims by 2030 to reduce the global malaria burden by 90% from 2015 levels. Non-falciparum malaria species have notably different biology than *P. falciparum* and are not necessarily controlled by the same measures, due to the potential for transmission by different vectors, differing seasonality[10], potentially earlier gametocytogenesis[11–13], presence of hypnozoite stages/persistent infection[12,14], generally lower levels of parasite carriage/density[15,16], and greater asymptomatic infection and transmission[10].

*P. vivax* is the most widely distributed human malaria species globally[17]. The strength of Duffy selection with rapid sweep across SSA is testament to the morbidity and mortality that *P. vivax* has caused in the past[18]. Although generally less severe than *P. falciparum*[19], it can be more difficult to control and treat due to the presence of the relapsing hypnozoite stage in the liver[20]. Outside of Africa, e.g., in the Western Pacific, *P. vivax* has increased as *P. falciparum* control has succeeded[1]. The prevailing dogma for decades was that *P. vivax* was largely absent from Africa due to high prevalence of the Duffy-negative trait, which is widespread in most SSA populations[21]. However, recent evidence across Africa indicates increased *P. vivax* detection in Duffy-negative patients[22–24]. Nonetheless, it is still much less prevalent than *P. falciparum* and the clinical significance and management strategies of such infections are still unknown.

*P. malariae* and *P. ovale* spp. are much less studied than either *P. falciparum* or *P. vivax*. They have both been detected throughout Africa, but usually at much lower prevalence than *P. falciparum*[25]. Both are frequently detected in co-infections with *P. falciparum* rather than as mono-infections[15,25–28], although exceptions have been noted[29]. *P. ovale* spp. infections are generally found at low-density, while *P. malariae infections* can cause higher parasitemia[15]. *P. ovale* spp. have relapsing stages like *P. vivax*, which likely means that their control and elimination will be more difficult, as is true of *P. vivax*[12]. *P. malariae* is thought to be more prevalent than *P. ovale*, particularly in West Africa[5,6,14,15]. It does not have a liver stage, but can cause long-term persistent infection[14].

While there are data to suggest that non-falciparum malaria is widespread in sub-Saharan Africa[4,26,28,30], and that these prevalences may be increasing[3,5,31], rapid diagnostic tests (RDTs) currently deployed in Africa are better at detecting falciparum malaria than other species[32,33]. Given the preponderance of both low-density infections and multiple species co-infections with non-falciparum malaria species, as well as the lower likelihood of severe disease, these species are less likely to be detected during surveillance activities or at the point-of-care. While molecular diagnostics such as qPCR are readily capable of detecting both falciparum and non-falciparum *Plasmodium* spp., these tools are highly technical and resource-prohibitive, so are rarely used outside of laboratory research activities.

Tanzania has the third highest malaria burden in the world, accounting for 4.1% of global malaria deaths in 2021[1]. Malaria transmission in Tanzania is predominantly *P. falciparum* and highly heterogeneous by region and season (**Figure 1A**)[34]. Transmission is high and endemic along the coast and the Lake Zone, and low, unstable, and seasonal in large urban areas and highland regions, while the rest of the country is subject to moderate seasonal transmission[34].

**Figure 1.**
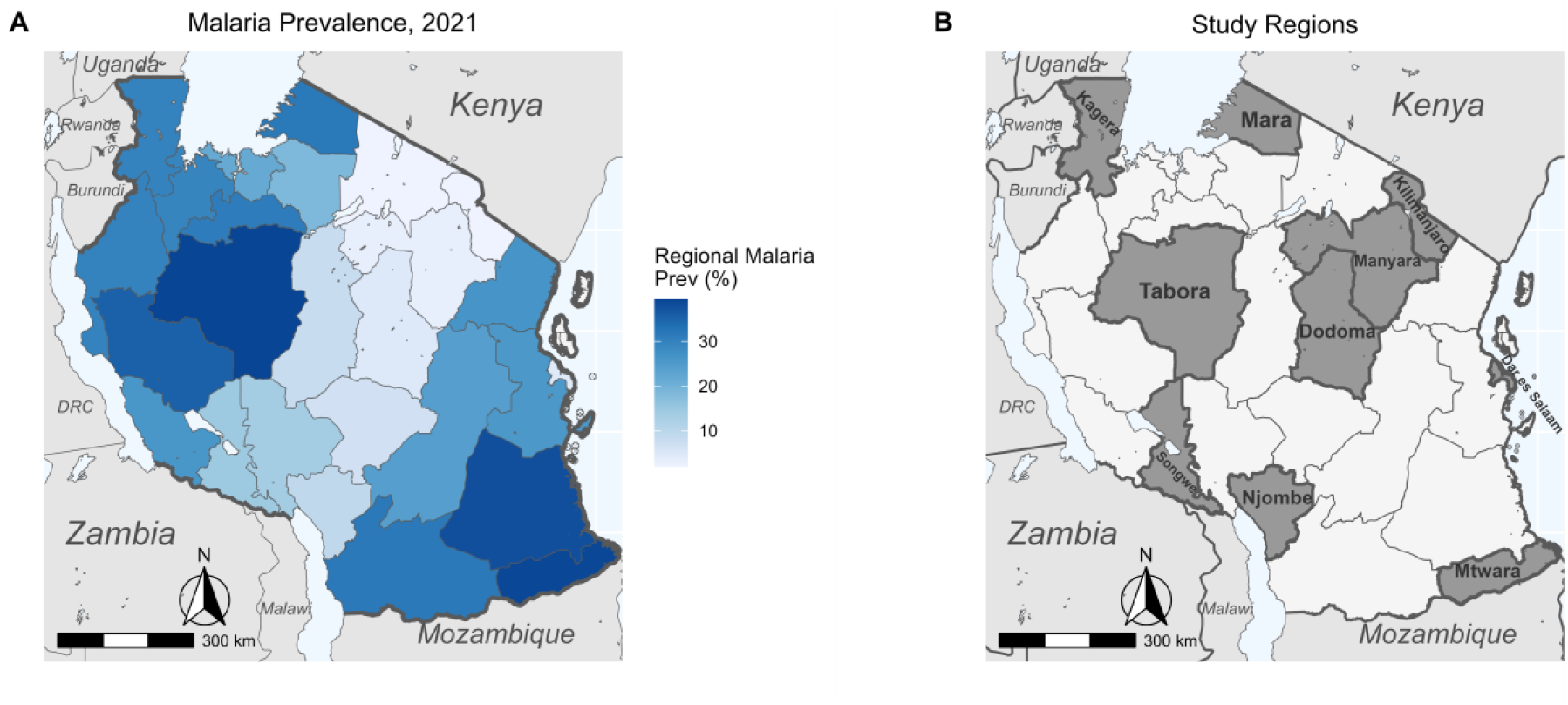
A) National map of regional malaria prevalence in 2021 as determined by a combination of blood slide positivity rates and *Pfhrp2*/LDH mRDT positivity rates. Data courtesy of National Malaria Control Programme, Dodoma. B) Map of Tanzania highlighting study regions.

Previous work has looked at prevalence of non-falciparum malaria among asymptomatic school children, finding surprisingly high rates of *P. ovale* spp. infection[8]. However, data from symptomatic cases for Tanzania is based primarily on sporadic convenience samples and does not provide broad geographic information about the distribution of non-falciparum malaria. To address the distribution of non-falciparum malaria in symptomatic individuals from all age groups, we conducted a survey of symptomatic malaria cases across ten different regions of Mainland Tanzania with varying transmission intensity. We present a molecular analysis of malaria species positivity rates within this national dataset.

## Methods

### Ethics

The Molecular Surveillance of Malaria in Tanzania (MSMT) study protocol was approved by the Tanzanian Medical Research Coordinating Committee (MRCC) of the National Institute for Medical Research (NIMR) and involved approved standard procedures for informed consent and sample deidentification. Additional details are described elsewhere[35]. Deidentified samples were considered non-human subjects’ research at the University of North Carolina and Brown University. In addition, publicly available aggregate data on malaria prevalence from all regions of Tanzania was provided by the National Malaria Control Programme (NMCP) for 2021 in order to map regional malaria prevalence.

### Sample collection

A random subset of 3,284 samples were drawn from 12,845 samples collected during the MSMT project[35] in 2021. Briefly, samples were collected in the MSMT *Plasmodium falciparum* histidine-rich protein 2 and 3 (*Pfhrp2/3*) gene deletion survey in ten regions: Dar es Salaam, Dodoma, Kagera, Kilimanjaro, Manyara, Mara, Mtwara, Njombe, Songwe, and Tabora[35], with additional samples collected in these regions for population genetic studies of malaria parasites. Additional details on study site selection are given in Rogier *et al*.[35]. Surveys were conducted according to the WHO protocol[36] and included ten regions (**Figure 1B**) which are distributed across mainland Tanzania with variable malaria transmission levels. Regions were assigned to four transmission strata ranging from very low to high based on NMCP data from 2020[34] (**Table 1**).

**Table 1.** Summary of study regions and number of samples analyzed.

| **Region** | **Transmission Strata** | **Samples** |
| --- | --- | --- |
| Kilimanjaro | Very Low | 897 |
| Manyara | Very Low | 223 |
| Njombe | Very Low | 211 |
| **Subtotal** | **Very Low** | **1,331** |
| Dar es Salaam | Low | 213 |
| Dodoma | Low | 274 |
| Songwe | Low | 145 |
| **Subtotal** | **Low** | **632** |
| Mara | Moderate | 335 |
| Tabora | Moderate | 267 |
| **Subtotal** | **Moderate** | **602** |
| Kagera | High | 417 |
| Mtwara | High | 302 |
| **Subtotal** | **High** | **719** |
| **Overall** | **All** | **3,284** |

Dried blood spots (DBS) were collected from patients with malaria-like symptoms. Most individuals (n = 3,234) were tested with a standard Pfhrp2/ pan-*Plasmodium* lactate dehydrogenase (pLDH) mRDT while some (n = 1,942) were also administered a PfLDH-based mRDT[35]. Samples were considered malaria-positive if any mRDT was positive.

### Molecular speciation assays

DNA from three 6mm DBS punches[37], representing approximately 25 μl of blood[8] was extracted using Chelex into a final usable volume of approximately 100 μl. Quantitative real time PCR assays targeting the 18S ribosomal subunit were performed according to published protocols (Supplemental Table 1)[28]. A separate qPCR assay was run for each species: *P. falciparum, P. malariae, P. ovale* spp. (detecting both *P. o. curtisi* and *P. o. wallikeri*), and *P. vivax*. Detection and parasitemia quantification were based on standard curves generated using dilutions of plasmids from MR4 (MRA-177, MRA-178, MRA-179, MRA-180; BEI Resources, Manassas, VA). Plasmids were quantified using a Qubit fluorometer, then normalized to a standard concentration of 0.1 ng/μL before serial dilutions. Three ten-fold dilution concentrations (0.001 ng/μL, 0.0001 ng/μL, 0.00001 ng/μL) were used for qPCR standard curves. Semi-quantitative parasitemia was estimated based on the assumption of six 18S rRNA gene copies per parasite genome[28], then multiplied by four to account for the dilution of eluted DNA relative to initial blood volume[8]. *P. malariae* assays used 42 cycles, while all other assays used 45 cycles to detect low-density infections[28]. This approach has been previously validated as highly sensitive and specific[28,38].

### Statistical analysis

Positivity rates were calculated for each region, as the data are biased toward individuals with both clinical symptoms and mRDT positivity and therefore do not represent true prevalence inclusive of asymptomatic infections. Regional-level maps of positivity rate for each species were created using the R package *sf* (version 1.0-9) based on shape files available from GADM.org and naturalearthdata.com accessed via the R package *rnaturalearth* (version 0.3.2)[39].

Variation in species-specific positivity rates by region, transmission strata, individuals’ age, and age group (young children <5 years, school-aged children 5-16 years[8,40], and adults >16 years) was assessed for significance with generalized linear models or ANOVA, as appropriate, in R. Given there is no consensus for an age range for school aged children, we used the age range of the 2017 Tanzania National School Children Survey[8,40].

## Results

### Study Population

Within this subset of 3,284 patients, the median age was 12 (IQR 3–28) years with a range of 6 months to 87 years. Children (≤16 years old) constituted 56.4% of participants (n = 1,853), while adults (>16 years old) constituted the remaining 43.6% (n = 1,431). Young children (<5 years old) comprised 58.3% (n = 1,081) of the child participants, while the remaining 41.7% (n = 772) were school-aged (5 – 16 years old). Gender identifications were available for 3,196 participants and female-skewed, with 1,754 female (54.9%) and 1,442 male participants (45.1%). Sample sizes for each region are given in **Table 1**. 53.7% (n = 1,763) of sampled individuals tested positive by any mRDT.

### qPCR Positivity by Species

Across all individuals sampled, *P. falciparum* was detected in 56.8% (n = 1,865, 95% CI: 55.1% – 58.5%), *P. malariae* in 1.7% (n = 55, 95% CI: 1.3% – 2.2%), *P. ovale* spp. in 5.0% (n = 163, 95% CI: 4.3% – 5.8%), and *P. vivax* in 0.09% (n = 3, 95% CI: 0.02% – 0.2%).

Mixed-species infections were common (**Table 2**). The majority of *P. malariae* infections were mixed with *P. falciparum* (56.4%, n = 31), although *P. malariae* mono-infections were also common (23.6%, n = 13), as were three-species infections with *P. malariae, P. falciparum* and *P. ovale* spp (18.2%, n = 10). The vast majority of *P. ovale* spp. infections were also mixed with *P. falciparum* (78.5%, n = 128, **Table 2**). As with *P. malariae*, single-species infections occurred and were more common than either three-species infections or mixed infection with *P. ovale* and *P. malariae* only. Of the three *P. vivax* infections detected, all were mixed with *P. falciparum* (**Table 2**). This small sample size precluded further statistical analyses.

**Table 2.** Infection composition proportions for non-falciparum infections.

| Infection Type | Count | Proportion |
| --- | --- | --- |
| *Pm* | 13 | 23.6% |
| *Pm/Pf* | 31 | 56.4% |
| *Pm/Po* | 1 | 1.8% |
| *Pm/Pf/Po* | 10 | 18.2% |
| All *Pm* Infections | 55 |  |
| *Po* | 24 | 14.7% |
| *Po/Pf* | 128 | 78.5% |
| *Pm/Po* | 1 | 0.6% |
| *Pm/Pf/Po* | 10 | 6.1% |
| All *Po* Infections | 163 |  |
| *Pv* | 0 | 0% |
| *Pv/Pf* | 3 | 100% |
| All *Pv* Infections | 3 |  |

\*Sum of proportions may not equal 100% due to rounding.

### Parasite Density

Parasitemia as determined by semi-quantitative qPCR based on standard dilutions of plasmid varied significantly (ANOVA p < 0.001) by species, with *P. falciparum* parasitemia the highest at median (IQR) 1,226,000 p/μL (23,280 – 13,090,000 p/μL), followed by *P. malariae* at 389,920 p/μL (19,364 – 2,147,000 p/μL), and *P. ovale* spp. having the lowest median parasitemia at 3,970 p/μL (396 – 54,000 p/μL). A Tukey post hoc test detected a significant difference in parasitemia (p < 0.001) between *P. falciparum* and *P. ovale* spp., but not between the other species pairs. Parasitemia did not differ significantly between single and mixed-species infections (Supplemental Figure 1). The three *P. vivax* infections (parasitemia of 70, 4,704, and 61,400 p/μL) were not analyzed.

### Variability by Region and Transmission Strata

*P. falciparum* was detected in every region sampled (**Figure 2A**) with variable positivity rates (**Table 3**). *P. malariae* and *P. ovale* spp. were detected in every region except for Dar es Salaam (**Figure 2B, 2C**). *P. vivax* was only detected in Kilimanjaro region (**Figure 2D**).

**Table 3.** Positivity Rates by Region.

| Region | Pf Positive Samples | Pm Positive Samples | Po Positive Samples | Pv Positive Samples | Total Samples |
| --- | --- | --- | --- | --- | --- |
| Dar es Salaam | 161 (75.6%) | 0 (0%) | 0 (0%) | 0 (0%) | 213 |
| Dodoma | 174 (63.5%) | 5 (1.8%) | 15 (5.5%) | 0 (0%) | 274 |
| Kagera | 261 (62.6%) | 13 (3.1%) | 24 (5.8%) | 0 (0%) | 417 |
| Kilimanjaro | 209 (23.3%) | 2 (0.2%) | 4 (0.4%) | 3 (0.3%) | 897 |
| Manyara | 144 (64.6%) | 3 (1.4%) | 14 (6.3%) | 0 (0%) | 227 |
| Mara | 240 (71.6%) | 15 (4.5%) | 51 (15.2%) | 0 (0%) | 335 |
| Mtwara | 238 (78.8%) | 3 (1.0%) | 10 (3.3%) | 0 (0%) | 302 |
| Njombe | 124 (58.8%) | 6 (2.8%) | 15 (7.1%) | 0 (0%) | 211 |
| Songwe | 93 (64.1%) | 3 (2.1%) | 14 (9.7%) | 0 (0%) | 145 |
| Tabora | 221 (82.8%) | 5 (1.9%) | 16 (6.0%) | 0 (0%) | 267 |

**Figure 2.**
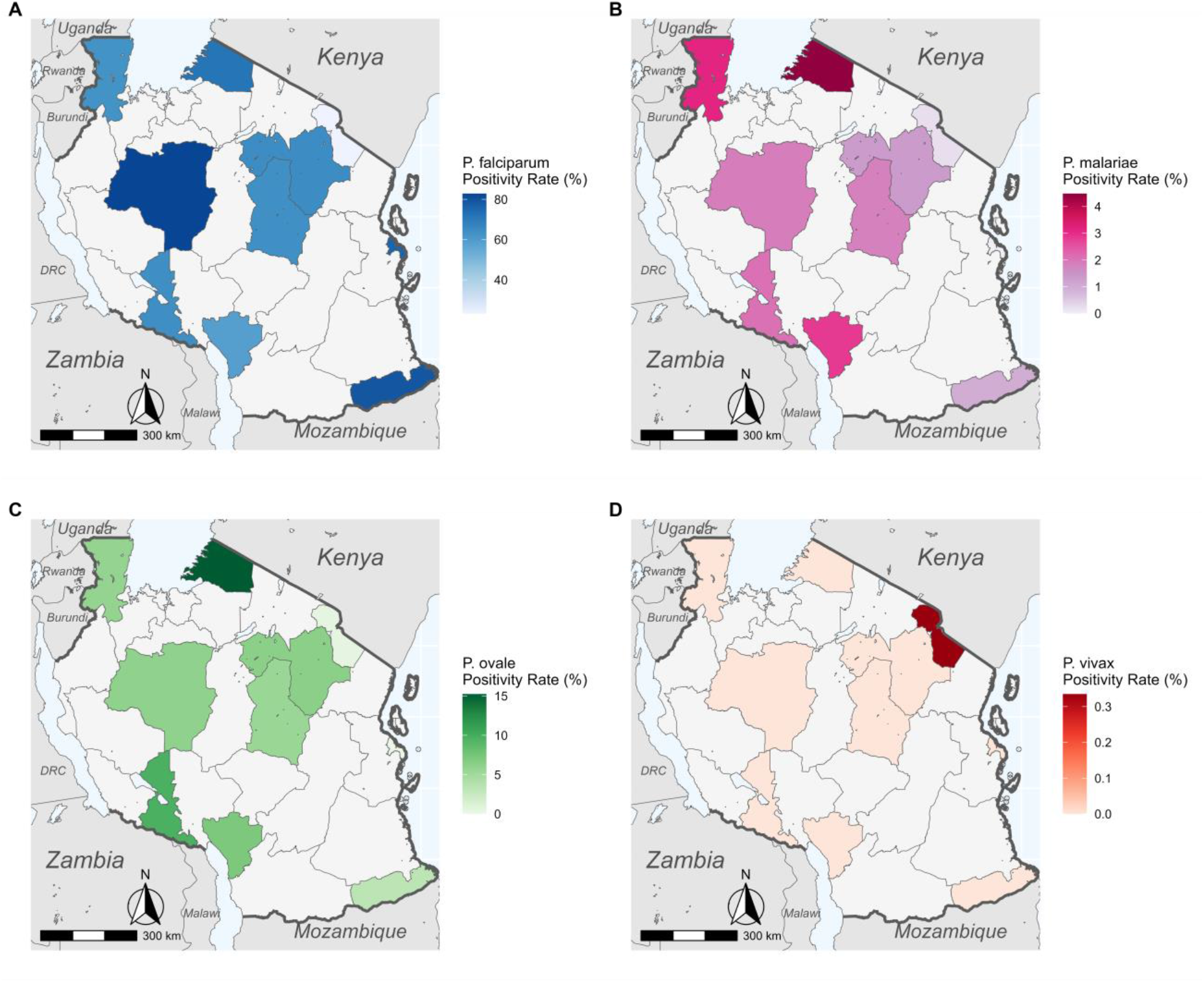
Species positivity rate maps across Tanzania for A) *P. falciparum*, B) *P. malariae*, C) *P. ovale* spp., *and* D) *P. vivax*. Shaded regions represent only positivity rates among health facility samples and should not be assumed to represent true regional prevalences.

*P. falciparum* was most abundant in Tabora (82.8%, n = 221/267, **Table 3**) and Mtwara (78.8%, n = 238/302), while it was least abundant in Kilimanjaro (23.3%, n = 209/897). *P. malariae* was most abundant in Mara (4.5%, n =15/335) and Kagera (3.1%, n = 13/417) and least abundant in Kilimanjaro (0.2%, n = 2/897) and Mtwara (1.0%, n = 3/302). *P. ovale* spp. positivity rates surpassed 5% in seven regions, with the highest abundance in Mara (15.2%, n = 51/335), Songwe (9.7%, n = 14/145), and Njombe (7.1%, n = 15/211). It was rare in Kilimanjaro (0.4%, n = 4/897). In all ten regions, *P. ovale* positivity rates were higher than *P. malariae*. Positivity rates varied significantly by region for all three species (ANOVA p < 0.001). Overall, transmission strata had a highly significant (ANOVA p < 0.001) effect on the positivity rates of all three species. Tukey post hoc analysis between strata is shown in **Figure 3**.

**Figure 3.**
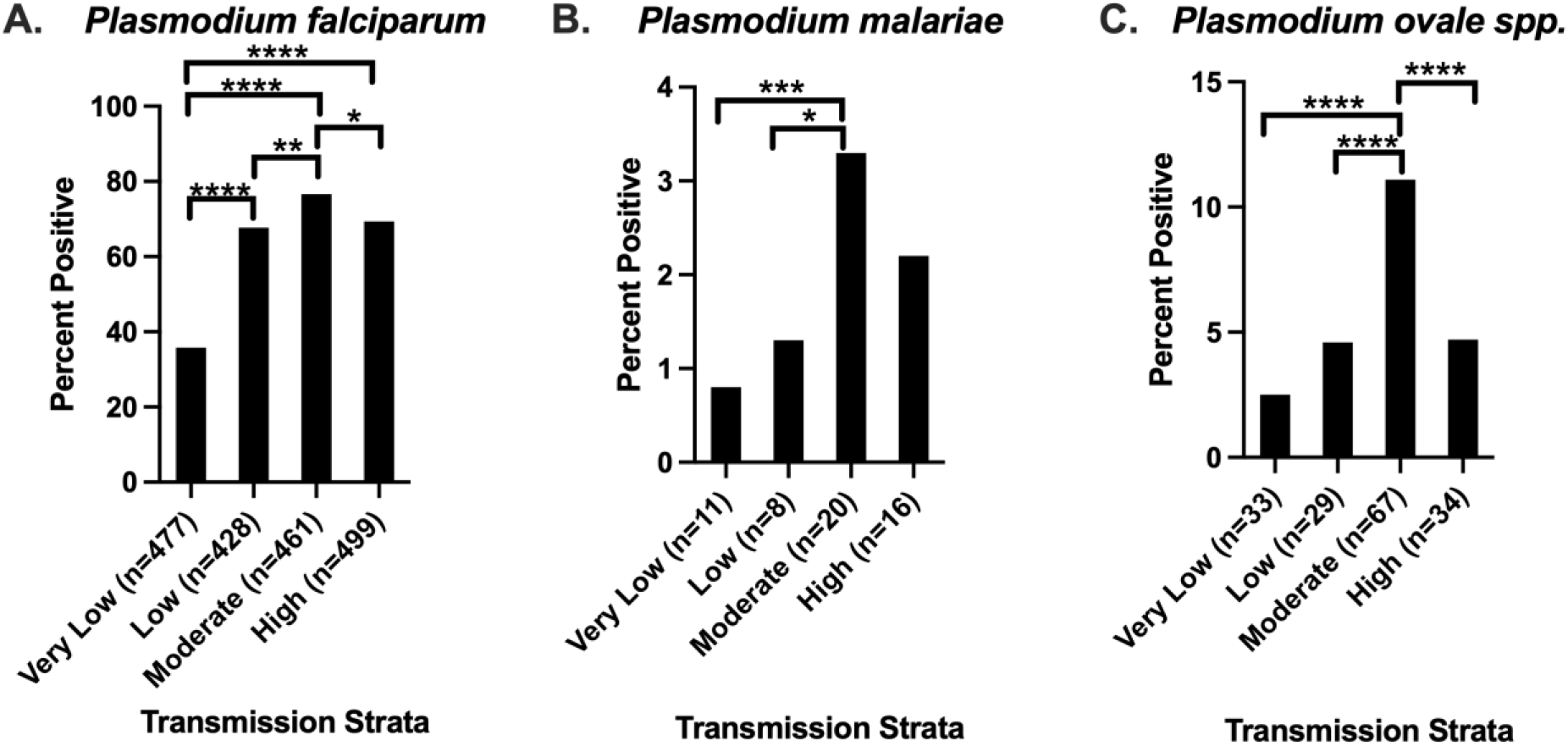
Tukey Analysis of Malaria Species Positivity Rate by Transmission Strata. A total of 1,331, 632, 602, and 719 samples were available in the very low, low, medium, and high transmission strata, respectively. The number of positive samples per strata for each species is shown in the X-axis labels. Level of significance is shown as follows: * p<0.05, ** p<0.01, *** p<0.001, or **** p<0.0001. Panel A shows *P. falciparum* parasite rates by strata. There were significant differences between most strata in pairwise comparison, therefore the only nonsignificant comparison is shown (NS). Panel B shows the *P. malariae* parasite rate by strata. Pairwise comparisons of strata with significant differences in parasite rate are shown (*). Panel C shows *P. ovale* spp. parasite rate by strata. Pairwise comparisons of strata with significant differences in parasite rate are shown (*).

Transmission strata had a significant effect on the likelihood of *P. falciparum* co-infection by ANOVA (p = 0.002) for *P. ovale* spp. A Tukey post hoc test indicated 24.6% (95% CI: 2.5% – 46.7%) fewer co-infections in very low vs. high transmission strata (p = 0.02) as well as 28.9% (95% CI: 0.97% – 48.1%) fewer co-infections in very low vs. moderate strata (p < 0.001). There was also a nearly significant (p = 0.06) reduction in co-infections between the very low and low transmission strata. By contrast, there was no significant effect on the likelihood of *P. malariae* co-infection with *P. falciparum*.

### Variability by Age Group

No significant difference in *P. malariae* prevalence by age in years was detected (GLM p = 0.24). In contrast to *P. malariae*, age significantly correlated with *P. ovale* spp. infection (GLM p = 0.001). Children had significantly more *P. ovale* spp. infections (6.5%, n = 120/1,853) than adults (3.0%, n = 43/1,431). When splitting participants into three age groups (**Figure 4**), overall significant correlations by ANOVA were observed with qPCR positivity for *P. falciparum* (p < 0.001), *P. malariae* (p = 0.004), and *P. ovale* spp. (p < 0.001) (**Figure 4**). For *P. falciparum*, a Tukey post hoc test indicated significant variation (p < 0.001) between young children (<5 years), school children (5 - 16 years), and adults (>16 years), with children more likely to be positive than adults and school children more likely to be positive than young children. School children were also more likely to be positive for *P. malariae* than either adults (p = 0.003) or young children (p = 0.03), but no significant differences were detected between young children and adults. For *P. ovale* spp., school children were again more likely to test positive than either adults (p < 0.001) or young children (p = 0.04), and young children were more likely to test positive than adults (p = 0.01).

**Figure 4.**
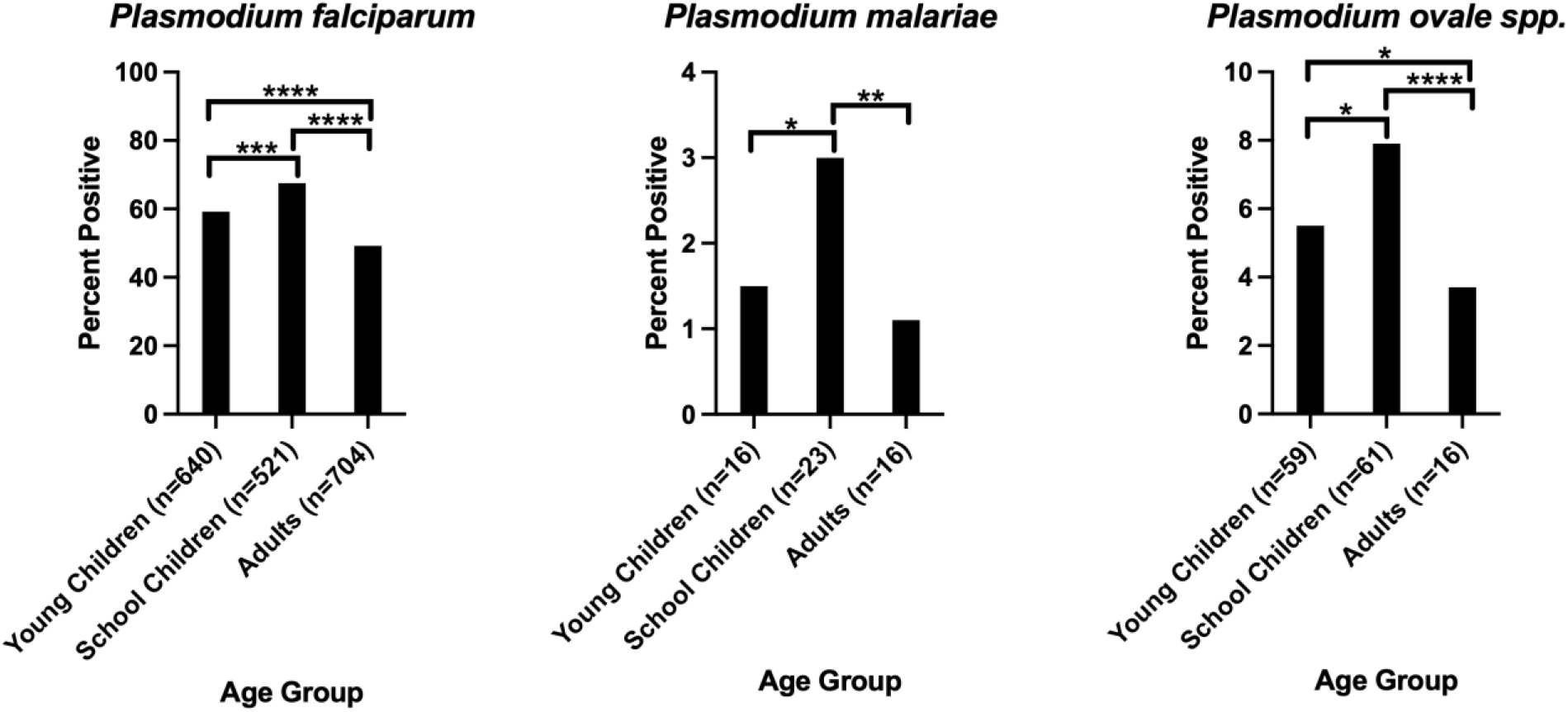
Tukey Analysis of Malaria Species Positivity Rate by Age Strata. A total of 1,081, 772, and 1,413 individuals were in the Young Children (<5 years), School Children (5-16 years) and Adult (>16 years) strata, respectively. The number of positive samples per age strata for each species is shown in the X-axis labels. Level of significance is shown as follows: * p<0.05, ** p<0.01, *** p<0.001, or **** p<0.0001. **Panel A** shows *P. falciparum* parasite rates by age strata. Pairwise comparisons of strata with significant differences in parasite rate are shown. All other pairwise comparisons were nonsignificant. **Panel B** shows the *P. malariae* parasite rate by strata. Pairwise comparisons of strata with significant differences in parasite rate are shown. All other pairwise comparisons were nonsignificant. **Panel C** shows *P. ovale* spp. parasite rate by strata. Pairwise comparisons of strata with significant differences in parasite rate are shown. All other pairwise comparisons were nonsignificant.

## Discussion

This study presents a comprehensive picture of the geographic distribution of different malaria species among symptomatic patients within Mainland Tanzania across all ages and in different transmission strata. It establishes a snapshot of the epidemiological landscape in 2021 that will enable effective longitudinal tracking of non-falciparum malaria. *Plasmodium falciparum* remains by far the most prevalent malaria species in Tanzania. Nonetheless, *P. malariae* and especially *P. ovale* spp. are much more abundant than previously acknowledged, with multiple regions approaching or surpassing (in the case of *P. ovale* spp.) 5% positivity rates. *P. vivax* is rarely detected and was found only in Kilimanjaro.

Our findings support a recent report of high rates of P. ovale infection among asymptomatic school aged children in 2017[8]. Direct comparison between the studies is not possible due to the significant methodological differences (e.g. clinic-based symptomatic versus school-based asymptomatic). However, we see predominately low density P. ovale infections that are over-represented in the school aged individuals in this study. Though the 2017 study identified *P. ovale* spp. primarily as mono-infections[8], we identified mostly mixed infections with *P. falciparum* (**Table 2**).This discrepancy is most likely explained by our inclusion of symptomatic patients in the study, as infection with *P. falciparum* likely led them to seek medical care, while school children with *P. ovale* spp. mono-infections are more likely to be asymptomatic[15]. Indeed, they constitute a major asymptomatic infectious reservoir[41–43].

The relatively high (> 5%) *P. ovale* spp. positivity rates in seven regions point to the necessity of active surveillance for these species, particularly given data from Tanzania and elsewhere showing increased incidence[3,5,7,31]. As was seen by the 2017 study[8], we found *P. ovale* spp. mono-infections to be more frequent in areas of low *P. falciparum* transmission, even though our samples came from symptomatic patients reporting to health facilities. The presence of these infections suggests that measures developed for *P. falciparum* will not completely control malaria in these regions. Since *P. ovale* spp. develops hypnozoites which can cause relapse, and which are not treated by standard schizonticidal regimens[12], its control is more complicated than that of *P. falciparum*. While major aspects of its biology, including major vectors, are poorly understood, transmission may be seasonal and asynchronous with *P. falciparum*[10]. Proactive monitoring will allow the NMCP and other decision makers to regularly monitor incidence and implement *P. ovale*-specific control measures as appropriate to support malaria control and elimination efforts.

While *P. malariae* was less abundant than *P. ovale* spp. in our study, it is nonetheless present throughout Tanzania and also warrants regular surveillance. We identified few significant differences in *P. malariae* positivity by transmission strata and no effect of transmission strata on likelihood of co-infection with *P. falciparum*, but these results may stem from small sample sizes (55 total *P. malariae* infections and 13 mono-infections). The relatively low *P. malariae* levels detected in our study are also consistent with those found in other surveys of Tanzania as well as Malawi and Western Kenya[8,28,44]. Taken together, these results suggest that *P. malariae* is less of a concern than *P. ovale* spp. in East Africa. However, transmission of *P. malariae*, like*P. ovale* spp., has been noted to increase as *P. falciparum* transmission reduces throughout East Africa[2,3,5,16]. If these trends continue and both species become more abundant, effective malaria control will increasingly hinge upon their surveillance.

While there was significant regional variation in *P. malariae* and *P. ovale* spp. abundance in our study, it did not consistently correspond with transmission strata defined by *P. falciparum* data. In both *P. malariae* and *P. ovale* spp., the moderate transmission strata had the highest positivity rate, largely stemming from the high rates of both in Mara region. While *P. malariae* was equally likely to be co-infected with *P. falciparum* in all transmission strata, *P. ovale* spp. was more likely to occur as a single-species mono-infection in the very low transmission strata.

Based on our data, *P. vivax* remains a minimal concern in Tanzania. However, as with the other species, its incidence could increase as *P. falciparum*’s decreases and like *P. ovale* spp., it would require specialized treatment and control measures. Other studies have also detected *P. vivax* at low levels in Kagera, Mara, and Mwanza[8,45], the coastal regions of Mtwara and Tanga[8], and on the Zanzibar archipelago[16,46]. The fact that we did not detect *P. vivax* in multiple regions is consistent with its characteristically low prevalence of ≤1%. Because of its low prevalence, more intensive and targeted surveillance will be necessary to understand the biology of *P. vivax* in Tanzania, but its clinical impact is likely minor compared to the other species.

Case-based surveillance pilots targeting malaria elimination have been implemented within the very low transmission regions of Kilimanjaro and Manyara, with the goal of expanding to other very low transmission regions[47]. While our results suggest that non-falciparum species are present at low prevalence within Kilimanjaro, and therefore unlikely to present major barriers to elimination in that region, Manyara has *P. ovale* spp. positivity rates > 5%, meaning that elimination efforts within that region will need to account for the specific surveillance, treatment, and control challenges presented by this species. Standardized treatment protocols currently employed in Tanzania do not incorporate “radical cure” with primaquine or tafenoquine to treat the liver-stage hypnozoites that cause relapses in *P. ovale* spp.[48]. As such, while ACT treatment may clear blood-stage parasites, relapses will continue to occur.

This study has several limitations. Given the sampling scheme of using only people presenting to the clinic with symptoms (whether they had mRDT-positive malaria or not) we are unable to determine true prevalence rates for each species and report parasite rate. This is due to the fact that mRDT-positive individuals are likely over-enrolled and the asymptomatic reservoir is not sampled. The addition of community surveys could help alleviate this. Parasitemia estimates are also relative given the use of plasmids as the control and a multi-copy gene for detection. Better controls are needed for accurate quantification of non-falciparum malaria using molecular methods. Lastly, while encompassing large regions of the country, the survey is not fully nationally representative and misses one crucial pre-elimination area, Zanzibar, where data on non-falciparum malaria may be very beneficial for elimination plans.

Non-falciparum malaria, particularly *P. ovale* spp. and *P. malariae*, is present throughout mainland Tanzania. In order to achieve effective control and elimination of malaria, it will be necessary to conduct further surveillance and research on these species. Ongoing research as part of MSMT will enable both longitudinal epidemiological study and genomic characterization of the non-falciparum malaria landscape in Tanzania, as nationwide samples from 2022 and 2023 will be available, allowing a fuller picture of the abundance of *P. malariae, P. ovale* spp., and *P. vivax* within Tanzania, as well as an indication of temporal trends that are not detectable in the work presented here. In addition, efforts to sequence high parasitemia *P. malariae* and *P. ovale* spp. samples collected in 2021 are ongoing and will grant us a deeper and more comprehensive picture of the Tanzanian populations of these species.

## Data Availability

National malaria transmission data was provided by the National Malaria Control Programme in Dodoma and is available by contacting NMCP. All other data is available upon reasonable request to the corresponding author.

## Acknowledgements

The authors wish to thank participants and parents/guardians of all children who took part in the surveillance. We acknowledge the contribution of the following project staff and other colleagues who participated in data collection and/or laboratory processing of samples: Raymond Kitengeso, Ezekiel Malecela, Muhidin Kassim, Athanas Mhina, August Nyaki, Juma Tupa, Anangisye Malabeja, Emmanuel Kessy, George Gesase, Tumaini Kamna, Grace Kanyankole, Oswald Osca, Richard Makono, Ildephonce Mathias, Godbless Msaki, Rashid Mtumba, Gasper Lugela, Gineson Nkya, Daniel Chale, Richard Malisa, Sawaya Msangi, Ally Idrisa, Francis Chambo, Kusa Mchaina, Neema Barua, Christian Msokame, Rogers Msangi, Salome Simba, Hatibu Athumani, Mwanaidi Mtui, Rehema Mtibusa, Jumaa Akida, Ambele Yatinga, and Tilaus Gustav. We also acknowledge the finance, administrative and logistic support team at NIMR: Christopher Masaka, Millen Meena, Beatrice Mwampeta, Gracia Sanga, Neema Manumbu, Halfan Mwanga, Arison Ekoni, Twalipo Mponzi, Pendael Nasary, Denis Byakuzana, Alfred Sezary, Emmanuel Mnzava, John Samwel, Daud Mjema, Seth Nguhu, Thomas Semdoe, Sadiki Yusuph, Alex Mwakibinga, Rodrick Ulomi and Andrea Kimboi. We are also grateful to the management of the National Institute for Medical Research, National Malaria Control Program and President’s Office-Regional Administration and Local Government (regional administrative secretaries of the 14 regions, and district officials, staff from all 100 HFs and Community Health Workers from the 4 community cross sectional regions). Technical and logistics support from the Bill and Melinda Gates Foundation team is highly appreciated. The following reagents were obtained through BEI Resources, NIAID, NIH: Diagnostic Plasmid Containing the Small Subunit Ribosomal RNA Gene (18S) from *Plasmodium falciparum*, MRA-177; *Plasmodium vivax*, MRA-178; *Plasmodium malariae*, MRA-179; and *Plasmodium ovale*, MRA-180, contributed by Peter A. Zimmerman.

---

Supplemental Table 1 – qPCR Reactions

(uploaded as separate Excel sheet)

**Supplemental Figure 1.**
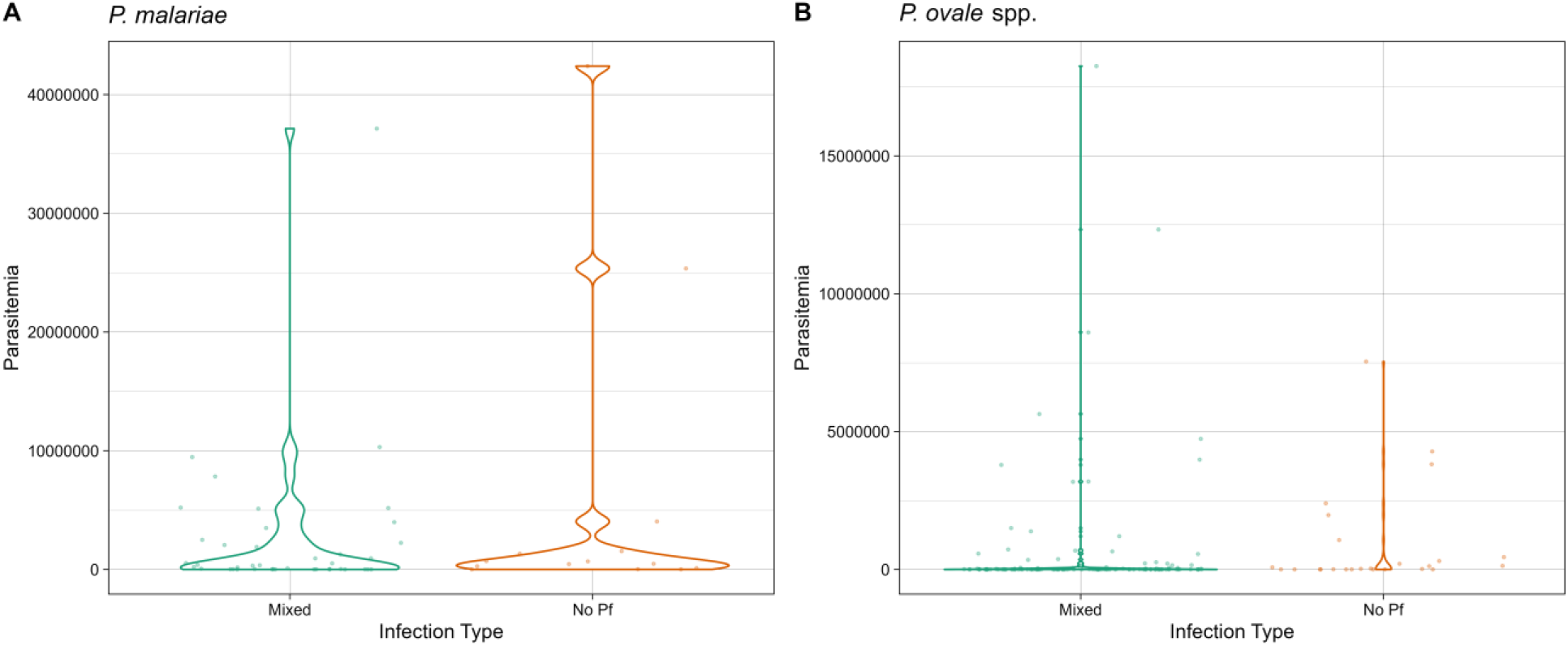
Parasitemia for Mixed Infections with *P. falciparum* vs. Single-Species Mono-Infections for (A) *P. malariae* and B) *P. ovale* spp. No significant difference was detected between mixed and mono-infections.

